# Emerging *Clostridioides difficile* strains belonging to PCR ribotype 955 in Serbia are distinct from metronidazole resistant RT955 outbreak isolates from the UK

**DOI:** 10.1101/2025.05.30.25328623

**Authors:** Predrag Stojanovic, Margriet Kraakman, Daan W. Notermans, James Groot, Céline Harmanus, Joffrey van Prehn, Mark Wilcox, Ed J. Kuijper, Wiep Klaas Smits, the Dutch National Expertise Centre for *Clostridioides difficile* group infections

## Abstract

End 2023, the UK Health Security Agency sent an alert about a new hypervirulent *Clostridioides difficile* PCR ribotype, ribotype 955 (RT955), causing slowly progressing infection clusters in hospitals in the Midlands. Between March 2018 and February 2022, CDI surveillance was performed in southern Serbia with centers providing medical services for approximately 750,000 inhabitants. Using the ECDC recommended protocol, clinical, epidemiological and microbiological data were collected. *C. difficile* RT955 was identified in 27 (7%) of 383 surveyed patients with CDI. Of 27 patients, 16 (59%) was older than 60 years and 19 (70%) were male. CDI was always associated with previous antibiotic therapy and had a hospital onset in 23 (85%) patients. The clinical presentaiton was milder than reported in UK and at 90 days follow up, no CDI related mortality was found. All sequenced strains belonged to multilocus sequence type (ST) 1 and were highly similar, with 0-1 alleles differences in a cgMLST analysis. The strains differed clearly from the UK RT955 outbreak strain by whole genome sequencing and phenotypic resistance to lincosamides and rifampicin. Interestingly, a high level erythromycin resistance was observed without a genetic resistance determinant. Both the UK and Serbian RT955 strains contained *gyrA*_p.T82I associated with resistance to fluoroquinolone antimicrobials and carried the P*nimB*^G^ promoter mutation, suggestive for haem-dependent metronidazole resistance. We conclude that *C. difficile* RT955 is present in Serbia since 2018, without presentation of large outbreaks. The Serbian RT955 strains differed clearly from a representative UK cluster strain, but shared its haem dependent metronidazole resistance.

## Introduction

Since 2003, *Clostridioides difficile* belonging to PCR ribotype (RT) 027 emerged and spread worldwide. Infections with RT027 were accompanied with higher mortality and morbidity and a more complicated course of the disease with frequent relapses and are for that reason sometimes referred to as hypervirulent (1). RT027 strains belong to multilocus sequence type (ST) 1, and phylogenetic clade 2. This clade also encompasses PCR ribotypes (such as 016, 036, 176, 181, 198, 244, and 251) that were later found and considered as “hypervirulent” but did not spread yet on a global scale (2, 3).

In response to these epidemiological developments, the European Centre for Disease Prevention and Control (ECDC) started the European *Clostridium difficile* Infection Surveillance Network (ECDIS-net), with a consortium composed of Universities from Leeds, Leiden, Wales, Berlin and the National Institute for Public Health and the Environment in the Netherlands (RIVM) to develop a European surveillance program. This protocol was subsequently tested, optimized and implemented (4). In 2018, a special program was added for EU candidate and potential candidate countries, including Serbia (IPA5 contract 4 within framework contract ECDC/2016/016

At the end of 2023, the UK Health Security Agency (UKHSA) issued an alert about a new hypervirulent RT955 causing slowly progressing clusters of infections in hospitals in the Midlands involving 50 patients with *C. difficile* infection (CDI) and data related to this PCR ribotype were presented at ESCMID Global conference in 2024 (5). Leiden retrospectively identified *C. difficile* RT955 strains among Serbian surveillance samples.

The aim of this descriptive report is to describe the emergence and spread of RT955 in a southern Serbia, with patients characteristics, clinical outcome, response to therapy and molecular findings.

## Methods

### Location

Between March 2018 and February 2022, CDI was surveyed in southern Serbia by the Institute of Public Health of Niš, Serbia, Centre for Microbiology (National Reference Laboratory for Anaerobic Infections – *C. difficile*). Several health care centers participate such as Community Health Center Niš, various hospitals, military hospitals and one clinical center in city Niš. These health care centers provide medical services for 750,000 individuals in the region. CDI is only tested by request of the physicians. Though a standardized surveillance protocol was lacking, the protocol of ECDC (6) was used as a basis for the diagnostics and case definitions.

### Definitions

CDI was defined as the occurrence of diarrhea (≥3 loose stools per day for at least 2 subsequent days) or the presence of a toxic megacolon and endoscopically diagnosed pseudomembranous colitis (PMC) combined with a positive test for toxin-producing *C. difficile*. Healthcare onset-CDI (HO-CDI) was defined as CDI in all patients who had onset of symptoms in a healthcare facility from 72 hours after admission until discharge. Community onset-CDI (CO-CDI) was defined as CDI in all patients who had onset of symptoms outside a healthcare facility at home. A recurrent CDI was defined as a new episode of diarrhoea occurring within 8 weeks after the day of onset of the previous CDI, with a positive microbiological diagnoss. The severity of CO-CDI was assessed as mild, moderate or severe using clinical characteristics as previously described (7).

### Microbiological analysis

**S**tool samples from patients with presumed CDI were analyzed by RIDA GENE *C. difficile* test (Real time multiplex PCR, R-Biopharm AG, Darmstadt, Germany) for the detection of *C. difficile* (16S rDNA) and *C. difficile* toxin A (*tcdA*) and toxin B (*tcdB*) genes. The presence of both free *C. difficile* toxins A and B in stool specimens was determined by the ELISA-RIDASCREEN *C. difficile* Toxin A/B (R - Biopharm AG, Darmstadt, Germany). Positive tested stool samples for CDI were inoculated on selective cycloserine-cefoxitin-fructose agar (CCFA) (Biomedics, Madrid, Spain) for *C. difficile* culture after an alcohol-shock treatment and incubated at 37 °C under anaerobic conditions for 48 h. Suspected *C. difficile* colonies were tested for the presence of glutamate dehydrogenase (GDH) using Rida®QUICK *C. difficile* GDH test (R - Biopharm AG, Darmstadt, Germany). Final identification of *C. difficile* was performed by the API system for anaerobic bacteria (API 20A bioMerieux, France). Isolates of *C. difficile* were send to Leiden University Medical Center (which hosts Dutch National Expertise Centre for *C. difficile* infections) for molecular characterization.

### Antimicrobial susceptibility testing

Antibiotic susceptibility testing was done both at the Institute of Public Health, Niš and at Leiden University Medical Center (LUMC). In Serbia, susceptibilities to five antibiotics were determined on Brucella Agar plates supplemented with haemin and vitamin K (Biomedics, Madrid, Spain) by E-test gradient strips (LIOFILCHEM ® srl, Italy), for vancomycin, tigecycline, fusidic acid, rifampicin, and moxifloxacin. Epidemiological cut-off values (ECOFF) of tested antibiotics were defined by European Committee on Antimicrobial Susceptibility Testing (EUCAST) guidelines (8). At the LUMC, isolates were retested for metronidazole susceptibility using a standardized and recommended agar dilution method (CLSI) on EUCAST-recommended Fastidious Anaerobe Agar supplemented with horse blood (FAA-HB) (9). In addition, 3 randomly selected RT955 strains were used for susceptibility testing to erythromycin, clindamycin and tetracyclin using Etest gradient strips (bioMérieux) on Brucella Blood Agar.

### Molecular characterization

*C. difficile* isolates were typed at the Dutch National Expertise Centre for *C. difficile* infections according to standard procedures by capillary PCR ribotyping (10). The presence of pCD-METRO was analyzed as described previously (11, 12). Whole genome sequencing was performed on an Illumina Novaseq 6000 platform and analyzed using the *C. difficile* cgMLST v2 and AMRFinderPlus routines implemented in SeqSphere+ 9.0.10 (Ridom GmbH, Germany). A minimal spanning tree (MST) was generated in SeqSphere+ with an MST cluster distance threshold of 6 (2). Mutations in the *nimB* gen and its promoter were analyzed by manually identifying the *nimB* gene and aligning the DNA sequence of the gene and its upstream region against reference sequences obtained for the historic RT027 strain CD196 (CD196_1331 in NC_013315.1) and the epidemic strain R20291 (the CDR20291_1308 (in NC_013316.1) in Geneious R10.2.6. (Biomatters Ltd). A SNP tree was generated based on 1833 SNPs in CSIPhylogeny 1.4 (13) using the genome sequence of strain CD196 as a reference; the resulting tree was rooted on strain CD196 (GBR-29-027 in our data). The tree was visualized in iTol (14) in rectangular mode ignoring branch lengths.

### Reuse of published WGS of *C. difficile*

Previously published genome sequences of *C. difficile* strains belonging to ST1 with frequently found PCR ribotypes (027, 176, 181) from various countries were included for comparison with the UK RT955 strain (GBR-30-955 in our analyses; corresponding to GenBank accession ERR12670107). A strain identified in Enterobase as RT955 (GBR-83-955 in our analyses), was also included. Most strains have been used in previous studies to compare PCR ribotyping with whole genome sequence typing and from an outbreak in Greece (2, 15)(Supplemental Table 1).

### Ethical approval

All procedures performed in studies involving human participants were in accordance with the ethical standards of the institutional and/or national research committee and with the 1964 Helsinki declaration and its later amendments or comparable ethical standards.

## Results

In total, 383 patients (290 healthcare onset [HO] and 93 community onset [CO] cases; 424 isolates) with CDI were included in the surveillance of which the majority derived from Clinical center Niš (284 patients) and Community Health Center Niš (93 patients). Six patients were diagnosed at the Military Hospital Nis. *C. difficile* RT 955 was identified in 28 of 424 isolates (7%), representing 27 of 383 patients (7 %). Four patients with CDI and RT 955 were also part of a previous report; SRB-02-955, SRB-05-955, SRB-09-955 and SRB-15-955 (16). Of the 27 patients with CDI due to RT955, 59% were older than 60 years and 70% were male (**Table 1**).

**Table 1.** Demographic data and clinical characteristics of Serbian patients with CDI and RT955

| Patients with CDI and RT 955 |  |
| --- | --- |
| N=27 |  |
| **Age** | 1 (4%) |
| Above 80 | 15 (56%) |
| Between 60 and 80 | 9 (33%) |
| Between 50 and 60 | 2 (7%) |
| Younger than 50 |  |
| **Male gender** | 19 (70%) |
| **Onset of CDI** |  |
| Hospital | 23 (85%) |
| Community | 4 (15%) |
| **Clinical presentation of CDI** |  |
| Mild | 11 (41%) |
| Moderate | 13 (48%) |
| Severe | 3 (11%) |
| **Previous use of antibiotics** |  |
| Cephalosporines | 20 (74%) |
| Fluoroquinolones | 7 (26%) |
| Macrolides | 6 (22%) |
| Combination of various antibiotic classes | 17 (63%) |
| **Concomitant disease** |  |
| COVID-19 | 16 (59%) |
| Gastrointestinal | 5 (19%) |
| Solid tumor | 3 (11%) |
| Chronic kidney disease | 2 (7%) |
| Cardiovascular disease | 1 (4%) |
| **Outcome after 90 days** |  |
| Recovered | 25 (93%) |
| All-cause mortality | 2 (7%) |
| Recurrence | 3 (11%) |
| **Treatment of CDI** |  |
| Only metronidazole | 6 (22%) |
| Only vancomycin | 11 (41%) |
| Only fidaxomicin | 1 (4%) |
| Metronidazole switch to vancomycin | 9 (33%) |

It was not possible to identify outbreaks of CDI due to RT955, since many hospitals were partially converted into COVID-19 centers with many movements of patients. CDI had a community onset in 4 (15%) patients and was always associated with previous antibiotic therapy, mostly including a cephalosporin in 20 patients (74%). Antibiotic combinations were used in 17 (63%) patients, mostly a combination of cephalosporines with fluoroquinolones (n=5) or with a macrolide (n=6). Of 27 patients, 16 (59%) had COVID-19 as concomitant disease whereas 5 patients (19%) had gastrointestinal disease, 3 (11%) a solid malignant tumor, 2 (7%) chronic kidney disease and 1 (4%) cardiovascular disease as underlying disease. One patient with HO-CDI presented as a recurrent episode of CDI. CDI was categorized as severe in 3 (11%) patients. At 90 days follow up, all patients recovered, 2 patients died but not related to CDI, and 3 (11%) developed a presumed recurrence. Of 15 patients treated with metronidazole, 9 (60%) patients switched to vancomycin.

Twenty-seven strains were available for WGS with cgMLST analysis using SeqSphere+. All sequenced Serbian strains were identified as ST1, CT 5259. Isolates were found to be highly related, with only 0-1 alleles differences in these analyses. Similar to other ST1 strains, the Serbian RT955 contained genes encoding toxin A (*tcdA*), toxin B (*tcdB*) and binary toxin (*cdtAB*), and carried a single base pair deletion at position 117 and an 18-bp deletion in the *tcdC* gene. A minimal spanning tree suggested that the Serbian strains are distinct from the UK RT955 cluster (**Figure 1**), as they fall in a different MST cluster.

**Figure 1.**
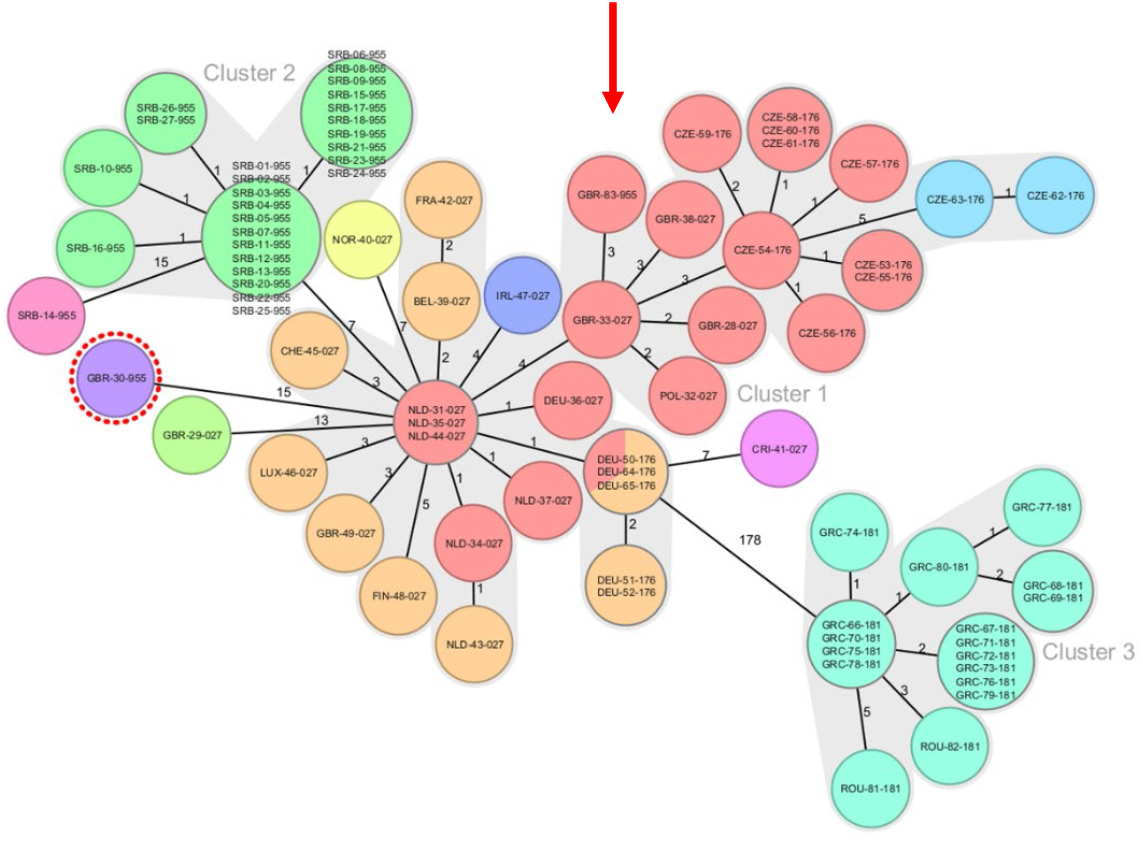
A minimal spanning tree based on cg MLST analysis with al Serbian RT955 strains, the UK RT955, and various RT027, RT176, and RT181 strains from various countries (2). Strain IDs include country of origin (first three letters per alpha-3 code), see Appendix Table 1. Each color represents a specific complex type. The numbers on the lines represent the number of allele differences between isolates. Three separate MST clusters are recognized, comprising RT027 and RT176 strains from various countries in Cluster 1, the majority of Serbian RT955 strains in Cluster 2 and RT181 strains from Romania and Greece in Cluster 3. The UK outbreak RT955 strain (GBR-30—955; purple with red dotted ring) shows 15 alleles difference from the RT027 cluster. The UK RT955 identified in Enterobase (GBR-83-955) is indicated with a red arrow

The phylogenetic tree based on whole genome SNP analysis using CSIPhylogeny (**Figure 2**) confirmed the high genomic relatedness of the Serbian RT955 strains (SRB), despite these mostly not being epidemiologically related. For 3 isolates we could identify an epidemiological link (i.e. the clinical data show a link in time and place) and the SNP analysis suggests that transmission may have occurred (Figure 1, strains SBR-25-955, SRB-26-955 and SRB-27-955). All 3 patients were diagnosed in October 2021 at Clinical Center Niš and were nursed in two adjacent rooms. Interestingly, two samples (SBR-15-955 and SBR-16-955) clearly differ from each other on a genomic level, but are derived from a single patient with a primary (SBR-15-955) and a recurrent CDI (SBR-16-955) after 4 weeks.

**Figure 2.**
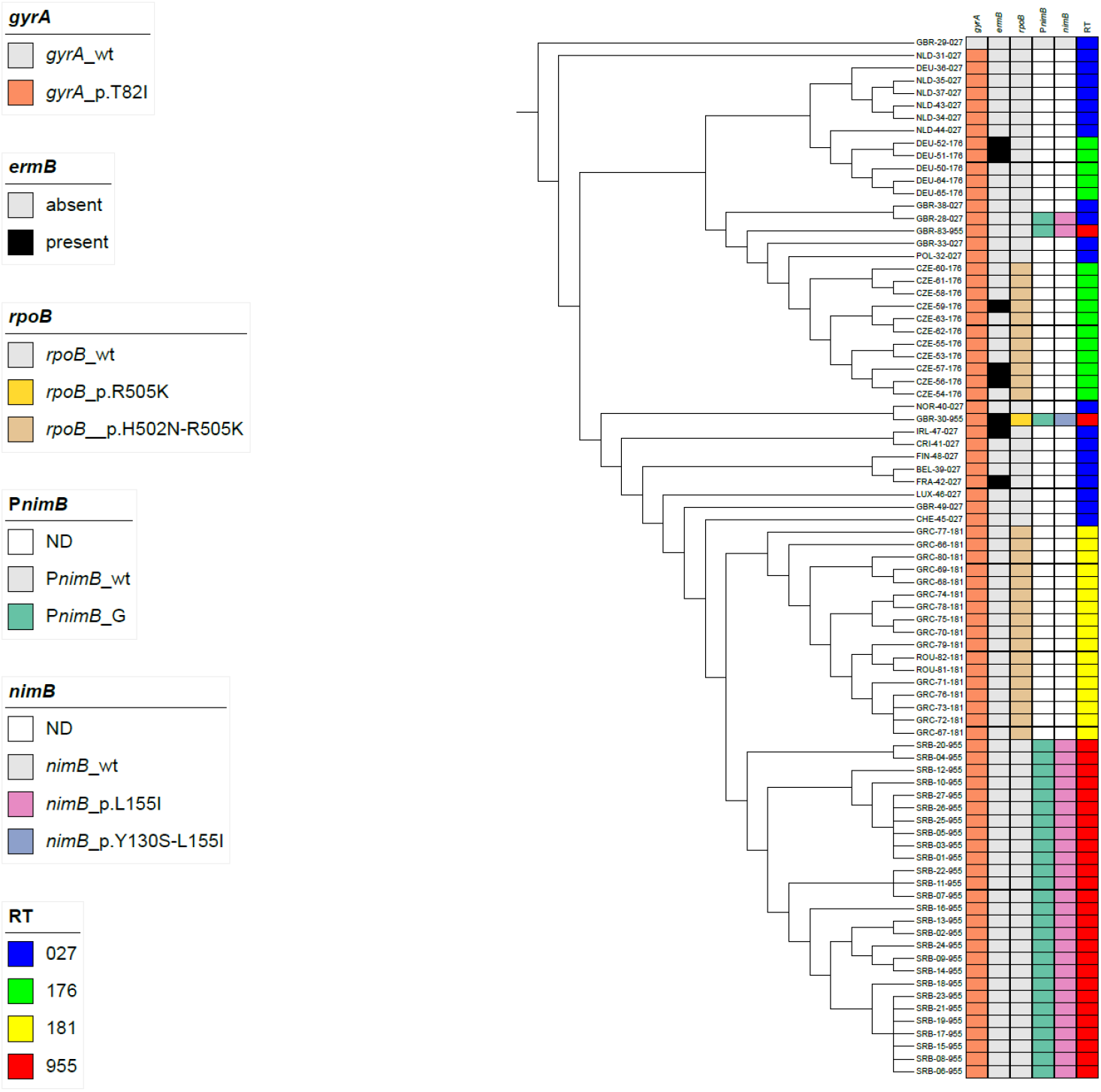
SNP-based phylogenetic tree of selected ST1 isolate. A SNP tree was generated based on 1833 SNPs in CSIPhylogeny 1.4 and visualised in iTOL v 6.9.2 (9). Strain IDs include country of origin (first three letters per alpha-3 code), see appendix Table 1. Historic and epidemic reference strains for RT027 (CD196 – GBR-29-027 and R20291 – GBR-028-027) are included, and CD196 was set as the root of the tree. Abbreviations: ND; not determined, *gyrA*; DNA gyrase A gene, *ermB*: erythromycin resistance gene, *rpoB*; RNA polymerase B gene, P*nimB*; promotor of nitroreductase gene, *nimB*; nitroreductase gene, RT; PCR ribotype.

An analysis of the antimicrobial resistance gene content (using AMRFinderPlus as implemented in SeqSphere+) shows that all RT955 isolates (both Serbian and UK) contain *gyrA*_p.T82I allele associated with fluoroquinolone resistance (**Figure 2**). In addition, manual analysis showed that all RT955 strains carried a T-to-G mutation in the promoter of of the *nimB* gene (P*nimB*^G^) implicated in medium-dependent metronidazole resistance (17)(Figure 2). We also note that – in comparison with the historic RT027 strain CD196 – all RT955 (both UK and Serbia) all carry a *nimB*_p.L155I mutation, but only the UK outbreak RT955 has an additional *nimB*_p.Y130S mutation. Other genomic resistance markers identified by AMRFinderPlus in the Serbian RT955 strains were for aph(2’’)-Ih (aminoglycoside resistance), and *blaCDD* (beta-lactam resistance).

Considering reported metronidazole resistance of the UK RT955 isolates, we investigated metronidazole susceptibility of the Serbian RT955 strains. Using the ECOFF criteria of EUCAST (8), Serbian RT955 strains were susceptible to metronidazole (n=20), vancomycin (n=27), tetracyclines (n=3), tigecyclin (n=27), fusidic acid (n=27) and rifampicin (n=27) by Etests on Brucella Blood Agar. Additional testing on FAA-HB, showed that all starins were found to be reduced susceptible to metronidazole with clearly elevated MICs (MIC 1-2 ug/mL). Interestingly, we also observed high erythromycin MICs (>256 ug/mL), but not clindamycin MICs, despite the fact that AMRFinderPlus did not identify erythromycin resistance determinants (18).

## Discussion

The UKHSA sent an alert of a new hypervirulent metronidazole resistant RT955 causing slowly progressing outbreaks in hospitals in Midlands in the period between September 2021 and December 2023, involving 50 patient with CDI. This raises the question whether RT955 has been circulating prior to these outbreaks and are present in other countries than the UK. Here we address these questions by showing that RT955 was present in Serbia prior to 2021. Subsequently, this RT was also found in Poland in at least three different hospitals (19). The Polish isolates in that study strongly resembled the UK outbreak strain. By contrast, we find that Serbian isolates differed in both a cgMLST and SNP analysis, despite belonging to the same ST (ST1). Importantly, the antibiotic susceptibility also differed with susceptibility of the Serbian strains to lincosamides and rifampicin, both phenotypically and genetically. The Serbian strains contained a similar gyrA_T82I mutation (associated with fluoroquinolones resistance) and a mutation in the promoter of the *nimB* gene (P*nimB*^G^) as did the UK outbreak strain. Phenotypical testing of antibiotic susceptibility, however, did not reveal metronidazole resistance on Brucella Blood agar (MIC= 0.5ug/mL). Notably, on FAA-HB we observed elevated MICs consistent with a heme-dependent mechanism of reduced susceptibility (MIC=1-2 mg/L) (9). Additional testing on different media with presumably higher levels of heme (e.g. chocolate agar with 1% polyvitex, PVX), we observed similar metronidazole MICs, demonstrating that lack of metronidazole resistance is likely not related to limiting haem concentrations (data not shown). We note, however, that the UK RT955 outbreak strain contains an additional *nimB*_p.Y130S mutation, of which the phenotypic consequences are unknown. Overall, our genomic and antimicrobial susceptibility data suggest that the Serbian isolates form a separate phylogroup.

Our data shows that RT955 isolates are common in Serbia beyond 2018. Of note, in 2013, Serbia participated in the European *Clostridium difficile* Infection Surveillance Network (ECDIS-Net) who launched a pilot study to enhance laboratory capacity and standardize surveillance for CDI, during which no *C. difficile* RT955 was identified; similarly, in a study performed during a 7-year period until 2018 in a different Serbian region, RT955 was not reported (20, 21). In Serbia, RT955 was also detected in 4% of the 93 samples included in a study to CO-CDI in the period from May 2018 to January 2022. It therefore appears that RT955 emerged in the last 5-6 years in Serbia, and has been commonly found.

Our epidemiological analyses are limited as a result of the impact of the COVID-19 pandemic. The Serbian RT955 described here were collected in the context of ECDC-supported CDI surveillance in southern Serbia. Though during the COVID-19 period this surveillance continued, the collection of clinical and epidemiological data to study outbreaks and transmission was hampered as many hospitals were converted into COVID centers. As a result it was not possible to determine the exact movement of all patients. In addition, infection prevention measures were reinforced at different stages in different hospitals to prevent spread of COVID-19. Nevertheless, spread and clustering of CDI with RT955 was suggested by our limited epidemiological data as well as our genomic analyses.

A few important observations on CDI due to the Serbian RT955 can be made. CDI affected mainly the elderly male patients and only 2 (7%) of 27 patients were found below the age of 50 years. CDI was mainly diagnosed as HO-CDI (85%) and always associated with previous antibiotic use. COVID-19 was reported as concomitant disease in 59% of the patients with CDI, and other well-known predisposing conditions for CDI were found in the remaining group. The high previous use of cephalosporines, fluoroquinolones and macrolides probably reflects the use of antibiotics for presumed bacterial pulmonary infections. The clinical presentation and outcome of CDI associated with RT955 differed from the UK experiences as reported at ESCMID Global 2024 (5). Most notably, between September 2021 and December 2023, 50 CDI cases of 48 patients were diagnosed in the UK, of which 11 died within 30 days. In the Serbian study, all-cause mortality was 7% and not attributed to CDI. Though only 11% of the Serbian patients reported a severe presentation of CDI, a treatment change from metronidazole to vancomycin was made in 50% of the patients who started with metronidazole. This may suggest a delayed response to metronidazole, potentially associated with the P*nimB*^G^ mutation, despite the fact that we did not observe metronidazole resistance under the conditions tested. Of note, elevated metronidazole MICs have been associated with clinical failure (22).

In summary, CDI with *C. difficile* RT955 is present in Serbia since 2018 with a milder clinical presentation than reported in the UK and without large outbreaks. The Serbian strain clearly differed from the UK reference strain at a genomic and phenotypic level.

Part of this work has been presented at 8^th^ International *C. difficile* symposium at Bled, 17-19 September 2024 (23).

## Data Availability

All data produced in the present work are contained in the manuscript.

## Data availability

Next generation sequencing data related to this manuscript are available through GenBank (BioProject PRJNA1225735) and detailed in Supplemental Table 1.

## Acknowledgements

We thank the administration of the Institute for Public Health Niš, Prof. Miodrag Stojanovic, Prof. Branislava Kocic and Prof. Dragan Bogdanovic for support with collection of clinical data. We also appreciate the support of dr. Karuna Vendrik with writing of the manuscript and dr. Igor Sidorov for uploading of the sequencing data

## Declaration of interest statement

The authors declare that they have no conflict of interest related to the work described here

## Funding statement

This work was supported by the Ministry of Education, Science and Technological Development of Serbia [Projects 172061; No. 451-03-65/2024-03/200113] and by the Faculty of Medicine. University of Nis, Serbia [Faculty of Medicine Niš, number 65. INT-MFN no. 65) to the prevalence and Incidence of hypervirulent *Clostridioides difficile* strains In Serbia. The support included the use of equipment and laboratories (without financial assistance).

---

**Supplemental Table 1.**
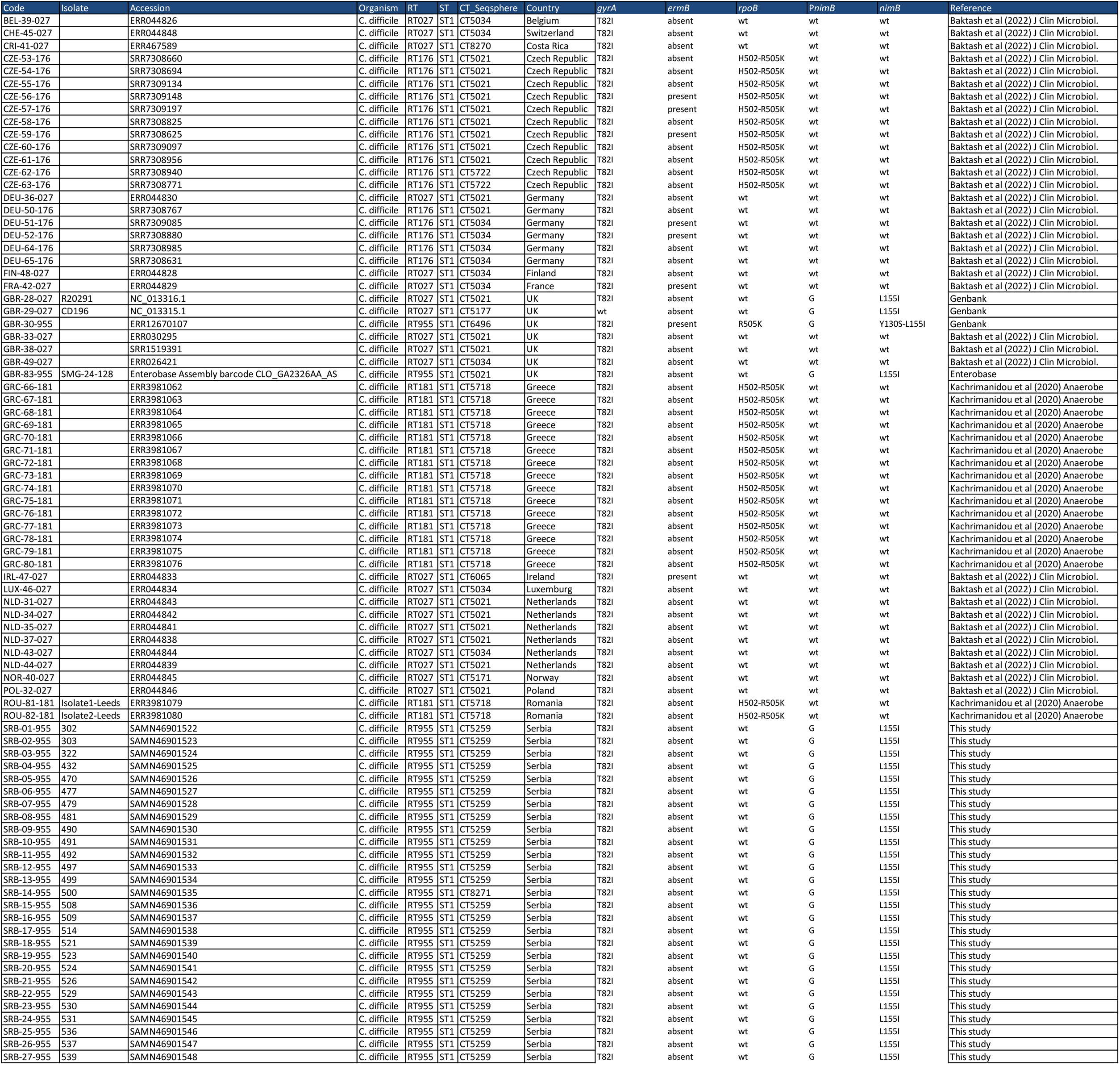

| Code | Isolate | Accession | Organism | RT | ST | CT_Seqsphere | Country | *gyrA* | *ermB* | *rpoB* | *PnimB* | *nimB* | Reference |
| --- | --- | --- | --- | --- | --- | --- | --- | --- | --- | --- | --- | --- | --- |
| BEL-39-027 |  | ERR044826 | C. difficile | RT027 | ST1 | CT5034 | Belgium | T82I | absent | wt | wt | wt | Baktash et al (2022) J Clin Microbiol. |
| CHE-45-027 |  | ERR044848 | C. difficile | RT027 | ST1 | CT5034 | Switzerland | T82I | absent | wt | wt | wt | Baktash et al (2022) J Clin Microbiol. |
| CRI-41-027 |  | ERR467589 | C. difficile | RT027 | ST1 | CT8270 | Costa Rica | T82I | absent | wt | wt | wt | Baktash et al (2022) J Clin Microbiol. |
| CZE-53-176 |  | SRR7308660 | C. difficile | RT176 | ST1 | CT5021 | Czech Republic | T82I | absent | H502-R505K | wt | wt | Baktash et al (2022) J Clin Microbiol. |
| CZE-54-176 |  | SRR7308694 | C. difficile | RT176 | ST1 | CT5021 | Czech Republic | T82I | absent | H502-R505K | wt | wt | Baktash et al (2022) J Clin Microbiol. |
| CZE-55-176 |  | SRR7309134 | C. difficile | RT176 | ST1 | CT5021 | Czech Republic | T82I | absent | H502-R505K | wt | wt | Baktash et al (2022) J Clin Microbiol. |
| CZE-56-176 |  | SRR7309148 | C. difficile | RT176 | ST1 | CT5021 | Czech Republic | T82I | present | H502-R505K | wt | wt | Baktash et al (2022) J Clin Microbiol. |
| CZE-57-176 |  | SRR7309197 | C. difficile | RT176 | ST1 | CT5021 | Czech Republic | T82I | present | H502-R505K | wt | wt | Baktash et al (2022) J Clin Microbiol. |
| CZE-58-176 |  | SRR7308825 | C. difficile | RT176 | ST1 | CT5021 | Czech Republic | T82I | absent | H502-R505K | wt | wt | Baktash et al (2022) J Clin Microbiol. |
| CZE-59-176 |  | SRR7308625 | C. difficile | RT176 | ST1 | CT5021 | Czech Republic | T82I | present | H502-R505K | wt | wt | Baktash et al (2022) J Clin Microbiol. |
| CZE-60-176 |  | SRR7309097 | C. difficile | RT176 | ST1 | CT5021 | Czech Republic | T82I | absent | H502-R505K | wt | wt | Baktash et al (2022) J Clin Microbiol. |
| CZE-61-176 |  | SRR7308956 | C. difficile | RT176 | ST1 | CT5021 | Czech Republic | T82I | absent | H502-R505K | wt | wt | Baktash et al (2022) J Clin Microbiol. |
| CZE-62-176 |  | SRR7308940 | C. difficile | RT176 | ST1 | CT5722 | Czech Republic | T82I | absent | H502-R505K | wt | wt | Baktash et al (2022) J Clin Microbiol. |
| CZE-63-176 |  | SRR7308771 | C. difficile | RT176 | ST1 | CT5722 | Czech Republic | T82I | absent | H502-R505K | wt | wt | Baktash et al (2022) J Clin Microbiol. |
| DEU-36-027 |  | ERR044830 | C. difficile | RT027 | ST1 | CT5021 | Germany | T82I | absent | wt | wt | wt | Baktash et al (2022) J Clin Microbiol. |
| DEU-50-176 |  | SRR7308767 | C. difficile | RT176 | ST1 | CT5021 | Germany | T82I | absent | wt | wt | wt | Baktash et al (2022) J Clin Microbiol. |
| DEU-51-176 |  | SRR7309085 | C. difficile | RT176 | ST1 | CT5034 | Germany | T82I | present | wt | wt | wt | Baktash et al (2022) J Clin Microbiol. |
| DEU-52-176 |  | SRR7308880 | C. difficile | RT176 | ST1 | CT5034 | Germany | T82I | present | wt | wt | wt | Baktash et al (2022) J Clin Microbiol. |
| DEU-64-176 |  | SRR7308985 | C. difficile | RT176 | ST1 | CT5034 | Germany | T82I | absent | wt | wt | wt | Baktash et al (2022) J Clin Microbiol. |
| DEU-65-176 |  | SRR7308631 | C. difficile | RT176 | ST1 | CT5034 | Germany | T82I | absent | wt | wt | wt | Baktash et al (2022) J Clin Microbiol. |
| FIN-48-027 |  | ERR044828 | C. difficile | RT027 | ST1 | CT5034 | Finland | T82I | absent | wt | wt | wt | Baktash et al (2022) J Clin Microbiol. |
| FRA-42-027 |  | ERR044829 | C. difficile | RT027 | ST1 | CT5034 | France | T82I | present | wt | wt | wt | Baktash et al (2022) J Clin Microbiol. |
| GBR-28-027 | R20291 | NC_013316.1 | C. difficile | RT027 | ST1 | CT5021 | UK | T82I | absent | wt | G | L155I | Genbank |
| GBR-29-027 | CD196 | NC_013315.1 | C. difficile | RT027 | ST1 | CT5177 | UK | wt | absent | wt | G | L155I | Genbank |
| GBR-30-955 |  | ERR12670107 | C. difficile | RT955 | ST1 | CT6496 | UK | T82I | present | R505K | G | Y130S-L155I | Genbank |
| GBR-33-027 |  | ERR030295 | C. difficile | RT027 | ST1 | CT5021 | UK | T82I | absent | wt | wt | wt | Baktash et al (2022) J Clin Microbiol. |
| GBR-38-027 |  | SRR1519391 | C. difficile | RT027 | ST1 | CT5021 | UK | T82I | absent | wt | wt | wt | Baktash et al (2022) J Clin Microbiol. |
| GBR-49-027 |  | ERR026421 | C. difficile | RT027 | ST1 | CT5034 | UK | T82I | absent | wt | wt | wt | Baktash et al (2022) J Clin Microbiol. |
| GBR-83-955 | SMG-24-128 | Enterobase Assembly barcode CLO_GA2326AA_AS | C. difficile | RT955 | ST1 | CT5021 | UK | T82I | absent | wt | G | L155I | Enterobase |
| GRC-66-181 |  | ERR3981062 | C. difficile | RT181 | ST1 | CT5718 | Greece | T82I | absent | H502-R505K | wt | wt | Kachrimanidou et al (2020) Anaerobe |
| GRC-67-181 |  | ERR3981063 | C. difficile | RT181 | ST1 | CT5718 | Greece | T82I | absent | H502-R505K | wt | wt | Kachrimanidou et al (2020) Anaerobe |
| GRC-68-181 |  | ERR3981064 | C. difficile | RT181 | ST1 | CT5718 | Greece | T82I | absent | H502-R505K | wt | wt | Kachrimanidou et al (2020) Anaerobe |
| GRC-69-181 |  | ERR3981065 | C. difficile | RT181 | ST1 | CT5718 | Greece | T82I | absent | H502-R505K | wt | wt | Kachrimanidou et al (2020) Anaerobe |
| GRC-70-181 |  | ERR3981066 | C. difficile | RT181 | ST1 | CT5718 | Greece | T82I | absent | H502-R505K | wt | wt | Kachrimanidou et al (2020) Anaerobe |
| GRC-71-181 |  | ERR3981067 | C. difficile | RT181 | ST1 | CT5718 | Greece | T82I | absent | H502-R505K | wt | wt | Kachrimanidou et al (2020) Anaerobe |
| GRC-72-181 |  | ERR3981068 | C. difficile | RT181 | ST1 | CT5718 | Greece | T82I | absent | H502-R505K | wt | wt | Kachrimanidou et al (2020) Anaerobe |
| GRC-73-181 |  | ERR3981069 | C. difficile | RT181 | ST1 | CT5718 | Greece | T82I | absent | H502-R505K | wt | wt | Kachrimanidou et al (2020) Anaerobe |
| GRC-74-181 |  | ERR3981070 | C. difficile | RT181 | ST1 | CT5718 | Greece | T82I | absent | H502-R505K | wt | wt | Kachrimanidou et al (2020) Anaerobe |
| GRC-75-181 |  | ERR3981071 | C. difficile | RT181 | ST1 | CT5718 | Greece | T82I | absent | H502-R505K | wt | wt | Kachrimanidou et al (2020) Anaerobe |
| GRC-76-181 |  | ERR3981072 | C. difficile | RT181 | ST1 | CT5718 | Greece | T82I | absent | H502-R505K | wt | wt | Kachrimanidou et al (2020) Anaerobe |
| GRC-77-181 |  | ERR3981073 | C. difficile | RT181 | ST1 | CT5718 | Greece | T82I | absent | H502-R505K | wt | wt | Kachrimanidou et al (2020) Anaerobe |
| GRC-78-181 |  | ERR3981074 | C. difficile | RT181 | ST1 | CT5718 | Greece | T82I | absent | H502-R505K | wt | wt | Kachrimanidou et al (2020) Anaerobe |
| GRC-79-181 |  | ERR3981075 | C. difficile | RT181 | ST1 | CT5718 | Greece | T82I | absent | H502-R505K | wt | wt | Kachrimanidou et al (2020) Anaerobe |
| GRC-80-181 |  | ERR3981076 | C. difficile | RT181 | ST1 | CT5718 | Greece | T82I | absent | H502-R505K | wt | wt | Kachrimanidou et al (2020) Anaerobe |
| IRL-47-027 |  | ERR044833 | C. difficile | RT027 | ST1 | CT6065 | Ireland | T82I | present | wt | wt | wt | Baktash et al (2022) J Clin Microbiol. |
| LUX-46-027 |  | ERR044834 | C. difficile | RT027 | ST1 | CT5034 | Luxemburg | T82I | absent | wt | wt | wt | Baktash et al (2022) J Clin Microbiol. |
| NLD-31-027 |  | ERR044843 | C. difficile | RT027 | ST1 | CT5021 | Netherlands | T82I | absent | wt | wt | wt | Baktash et al (2022) J Clin Microbiol. |
| NLD-34-027 |  | ERR044842 | C. difficile | RT027 | ST1 | CT5021 | Netherlands | T82I | absent | wt | wt | wt | Baktash et al (2022) J Clin Microbiol. |
| NLD-35-027 |  | ERR044841 | C. difficile | RT027 | ST1 | CT5021 | Netherlands | T82I | absent | wt | wt | wt | Baktash et al (2022) J Clin Microbiol. |
| NLD-37-027 |  | ERR044838 | C. difficile | RT027 | ST1 | CT5021 | Netherlands | T82I | absent | wt | wt | wt | Baktash et al (2022) J Clin Microbiol. |
| NLD-43-027 |  | ERR044844 | C. difficile | RT027 | ST1 | CT5034 | Netherlands | T82I | absent | wt | wt | wt | Baktash et al (2022) J Clin Microbiol. |
| NLD-44-027 |  | ERR044839 | C. difficile | RT027 | ST1 | CT5021 | Netherlands | T82I | absent | wt | wt | wt | Baktash et al (2022) J Clin Microbiol. |
| NOR-40-027 |  | ERR044845 | C. difficile | RT027 | ST1 | CT5171 | Norway | T82I | absent | wt | wt | wt | Baktash et al (2022) J Clin Microbiol. |
| POL-32-027 |  | ERR044846 | C. difficile | RT027 | ST1 | CT5021 | Poland | T82I | absent | wt | wt | wt | Baktash et al (2022) J Clin Microbiol. |
| ROU-81-181 | Isolate1-Leeds | ERR3981079 | C. difficile | RT181 | ST1 | CT5718 | Romania | T82I | absent | H502-R505K | wt | wt | Kachrimanidou et al (2020) Anaerobe |
| ROU-82-181 | Isolate2-Leeds | ERR3981080 | C. difficile | RT181 | ST1 | CT5718 | Romania | T82I | absent | H502-R505K | wt | wt | Kachrimanidou et al (2020) Anaerobe |
| SRB-01-955 | 302 | SAMN46901522 | C. difficile | RT955 | ST1 | CT5259 | Serbia | T82I | absent | wt | G | L155I | This study |
| SRB-02-955 | 303 | SAMN46901523 | C. difficile | RT955 | ST1 | CT5259 | Serbia | T82I | absent | wt | G | L155I | This study |
| SRB-03-955 | 322 | SAMN46901524 | C. difficile | RT955 | ST1 | CT5259 | Serbia | T82I | absent | wt | G | L155I | This study |
| SRB-04-955 | 432 | SAMN46901525 | C. difficile | RT955 | ST1 | CT5259 | Serbia | T82I | absent | wt | G | L155I | This study |
| SRB-05-955 | 470 | SAMN46901526 | C. difficile | RT955 | ST1 | CT5259 | Serbia | T82I | absent | wt | G | L155I | This study |
| SRB-06-955 | 477 | SAMN46901527 | C. difficile | RT955 | ST1 | CT5259 | Serbia | T82I | absent | wt | G | L155I | This study |
| SRB-07-955 | 479 | SAMN46901528 | C. difficile | RT955 | ST1 | CT5259 | Serbia | T82I | absent | wt | G | L155I | This study |
| SRB-08-955 | 481 | SAMN46901529 | C. difficile | RT955 | ST1 | CT5259 | Serbia | T82I | absent | wt | G | L155I | This study |
| SRB-09-955 | 490 | SAMN46901530 | C. difficile | RT955 | ST1 | CT5259 | Serbia | T82I | absent | wt | G | L155I | This study |
| SRB-10-955 | 491 | SAMN46901531 | C. difficile | RT955 | ST1 | CT5259 | Serbia | T82I | absent | wt | G | L155I | This study |
| SRB-11-955 | 492 | SAMN46901532 | C. difficile | RT955 | ST1 | CT5259 | Serbia | T82I | absent | wt | G | L155I | This study |
| SRB-12-955 | 497 | SAMN46901533 | C. difficile | RT955 | ST1 | CT5259 | Serbia | T82I | absent | wt | G | L155I | This study |
| SRB-13-955 | 499 | SAMN46901534 | C. difficile | RT955 | ST1 | CT5259 | Serbia | T82I | absent | wt | G | L155I | This study |
| SRB-14-955 | 500 | SAMN46901535 | C. difficile | RT955 | ST1 | CT8271 | Serbia | T82I | absent | wt | G | L155I | This study |
| SRB-15-955 | 508 | SAMN46901536 | C. difficile | RT955 | ST1 | CT5259 | Serbia | T82I | absent | wt | G | L155I | This study |
| SRB-16-955 | 509 | SAMN46901537 | C. difficile | RT955 | ST1 | CT5259 | Serbia | T82I | absent | wt | G | L155I | This study |
| SRB-17-955 | 514 | SAMN46901538 | C. difficile | RT955 | ST1 | CT5259 | Serbia | T82I | absent | wt | G | L155I | This study |
| SRB-18-955 | 521 | SAMN46901539 | C. difficile | RT955 | ST1 | CT5259 | Serbia | T82I | absent | wt | G | L155I | This study |
| SRB-19-955 | 523 | SAMN46901540 | C. difficile | RT955 | ST1 | CT5259 | Serbia | T82I | absent | wt | G | L155I | This study |
| SRB-20-955 | 524 | SAMN46901541 | C. difficile | RT955 | ST1 | CT5259 | Serbia | T82I | absent | wt | G | L155I | This study |
| SRB-21-955 | 526 | SAMN46901542 | C. difficile | RT955 | ST1 | CT5259 | Serbia | T82I | absent | wt | G | L155I | This study |
| SRB-22-955 | 529 | SAMN46901543 | C. difficile | RT955 | ST1 | CT5259 | Serbia | T82I | absent | wt | G | L155I | This study |
| SRB-23-955 | 530 | SAMN46901544 | C. difficile | RT955 | ST1 | CT5259 | Serbia | T82I | absent | wt | G | L155I | This study |
| SRB-24-955 | 531 | SAMN46901545 | C. difficile | RT955 | ST1 | CT5259 | Serbia | T82I | absent | wt | G | L155I | This study |
| SRB-25-955 | 536 | SAMN46901546 | C. difficile | RT955 | ST1 | CT5259 | Serbia | T82I | absent | wt | G | L155I | This study |
| SRB-26-955 | 537 | SAMN46901547 | C. difficile | RT955 | ST1 | CT5259 | Serbia | T82I | absent | wt | G | L155I | This study |
| SRB-27-955 | 539 | SAMN46901548 | C. difficile | RT955 | ST1 | CT5259 | Serbia | T82I | absent | wt | G | L155I | This study |

## Notes

### Competing Interest Statement

The authors have declared no competing interest.

### Author Declarations

All procedures performed in studies involving human participants were in accordance with the ethical standards of the institutional and/or national research committee and with the 1964 Helsinki declaration and its later amendments or comparable ethical standards. Ethical approval given by Etyckom Odboru (Ethics Committee), Institut Za Javno Zdravje Nis (Institute of Public Health Nis.

## References

1. He M, Miyajima F, Roberts P, Ellison L, Pickard DJ, Martin MJ, et al. Emergence and global spread of epidemic healthcare-associated Clostridium difficile. Nat Genet. 2013;45(1):109–13.

2. Baktash A, Corver J, Harmanus C, Smits WK, Fawley W, Wilcox MH, et al. Comparison of Whole-Genome Sequence-Based Methods and PCR Ribotyping for Subtyping of Clostridioides difficile. J Clin Microbiol. 2022;60(2):e0173721.

3. Smits WK, Lyras D, Lacy DB, Wilcox MH, Kuijper EJ. Clostridium difficile infection. Nat Rev Dis Primers. 2016;2:16020.

4. van Dorp SM, Kinross P, Gastmeier P, Behnke M, Kola A, Delmee M, et al. Standardised surveillance of Clostridium difficile infection in European acute care hospitals: a pilot study, 2013. Euro Surveill. 2016;21(29).

5. Puleston R, Roulston K, Morgan K, Hopkins S, Wilcox M, Fawley W, et al. O0430 - Emergence of new concerning ribotype of Clostridioides difficile (955). ESCMID Global; Barcelona, Spain 2024.

6. Control. ECfDPa. European surveillance of Clostridioides (Clostridium) difficile infections. Surveillance protocol version 2.4.. Stockholm: ECDC; 2019.

7. Ruuska T, Vesikari T. Rotavirus disease in Finnish children: use of numerical scores for clinical severity of diarrhoeal episodes. Scand J Infect Dis. 1990;22(3):259–67.

8. EUCAST. Clinical breakpoints 2024 [Available from: http://www.eucast.org/clinical_breakpoints/.

9. Freeman J, Sanders I, Harmanus C, Clark EV, Berry AM, Smits WK. Antimicrobial susceptibility testing of Clostridioides difficile: a dual-site study of three different media and three therapeutic antimicrobials. Clin Microbiol Infect. 2025.

10. Fawley WN, Knetsch CW, MacCannell DR, Harmanus C, Du T, Mulvey MR, et al. Development and validation of an internationally-standardized, high-resolution capillary gel-based electrophoresis PCR-ribotyping protocol for Clostridium difficile. PLoS One. 2015;10(2):e0118150.

11. Boekhoud IM, Hornung BVH, Sevilla E, Harmanus C, Bos-Sanders I, Terveer EM, et al. Plasmid-mediated metronidazole resistance in Clostridioides difficile. Nat Commun. 2020;11(1):598.

12. Smits WK, Harmanus C, Sanders I, Bry L, Blackwell GA, Ducarmon QR, et al. Sequence-Based Identification of Metronidazole-Resistant Clostridioides difficile Isolates. Emerg Infect Dis. 2022;28(11):2308–11.

13. Kaas RS, Leekitcharoenphon P, Aarestrup FM, Lund O. Solving the problem of comparing whole bacterial genomes across different sequencing platforms. PLoS One. 2014;9(8):e104984.

14. Letunic I, Bork P. Interactive Tree of Life (iTOL) v6: recent updates to the phylogenetic tree display and annotation tool. Nucleic Acids Res. 2024;52(W1):W78–W82.

15. Kachrimanidou M, Baktash A, Metallidis S, Tsachouridou O, Netsika F, Dimoglou D, et al. An outbreak of Clostridioides difficile infections due to a 027-like PCR ribotype 181 in a rehabilitation centre: Epidemiological and microbiological characteristics. Anaerobe. 2020;65:102252.

16. Stojanovic P, Harmanus C, Kuijper EJ. Community-onset Clostridioides difficile infection in south Serbia. Anaerobe. 2023;79:102669.

17. Olaitan AO, Dureja C, Youngblom MA, Topf MA, Shen WJ, Gonzales-Luna AJ, et al. Decoding a cryptic mechanism of metronidazole resistance among globally disseminated fluoroquinolone-resistant Clostridioides difficile. Nat Commun. 2023;14(1):4130.

18. Spigaglia P, Mastrantonio P, Barbanti F. Antibiotic Resistances of Clostridioides difficile. Adv Exp Med Biol. 2024;1435:169–98.

19. Zikova J, Szarek K, Kabała M, Wultańska D, Frankowska N, Iwanicki K, et al. The newly emerging metronidazole-resistant Clostridioides difficile PCR ribotype 955 has been identified in Poland in 2021–2023 but not in the Czech Republic and Slovakia. Euro Surveill. 2025;In press.

20. Jovanovic M, van Dorp SM, Drakulovic M, Papic D, Pavic S, Jovanovic S, et al. A pilot study in Serbia by European Clostridium difficile Infection Surveillance Network. Acta Microbiol Immunol Hung. 2019;67(1):42–8.

21. Predrag S, Kuijper EJ, Nikola S, Vendrik KEW, Niko R. Recurrent community-acquired Clostridium(Clostridioides)difficile infection in Serbianchildren. Eur J Clin Microbiol Infect Dis. 2020;39(3):509–16.

22. Gonzales-Luna AJ, Olaitan AO, Shen WJ, Deshpande A, Carlson TJ, Dotson KM, et al. Reduced Susceptibility to Metronidazole Is Associated With Initial Clinical Failure in Clostridioides difficile Infection. Open Forum Infect Dis. 2021;8(8):ofab365.

23. Predrag, S., Margriet, K., Joffrey, V. P., James, G., David, E.,Mark, W., Klaas, S, W. Kuijper. Clostridioides Difficile strains belonging to Ribotype 955, isolated during the Covid-19 pandemic in Southern Serbia, are genomically distinct from Ribotype 955 outbreak isolates from The Uk, 8th INTERNATIONAL C. difficile SYMPOSIUM, 17-19 September 2024 (Poster 49.

